# Neural Responsivity to Unexpected Stimuli Moderates a Developmental Pathway to Anxiety: A Community-Sample Replication in Early Infancy and Childhood

**DOI:** 10.64898/2026.09.17.26363348

**Authors:** Jiayin Xing, Christine Eun, Santiago Morales, Daniel S. Pine, Nathan A. Fox, Courtney A. Filippi

**Affiliations:** Department of Child and Adolescent Psychiatry, New York University, New York, NY, USA; Department of Psychology, University of Southern California, Los Angeles, CA, USA; Emotion and Development Branch, National Institute of Mental Health, Bethesda, MD, USA; Department of Human Development and Quantitative Methodology, University of Maryland, College Park, MD, USA

**Keywords:** mismatch response, negative reactivity, behavioral inhibition, infancy, anxiety

## Abstract

**Background:** Temperamental risk, including negative reactivity (NR) and behavioral inhibition (BI), represents a robust early pathway to anxiety. Neural responsivity to unexpected sensory stimuli, indexed by the mismatch response (MMR), has been shown to moderate associations linking early temperament to later anxiety. This study extends prior work by testing whether MMR measured as early as 4 months, in a community sample, moderate the developmental pathways from NR to BI and from BI to childhood anxiety.

**Methods:** Participants included 147 infants from the Origins of Infant Temperament study. NR and EEG were assessed at 4 months, with EEG recorded during an auditory oddball paradigm. BI was observationally measured at 14 months, and anxiety symptoms were assessed at 30 months and 4–6 years using the Child Behavior Checklist. Structural equation modeling with full-information maximum likelihood tested moderation effects of MMR on NR to BI and BI to anxiety pathways.

**Results:** MMR significantly moderated both pathways. More negative MMR at 4 months were associated with the risk pathways from high NR to high BI and from high BI to high anxiety at 30 months. No significant associations were observed at more positive MMR levels. Anxiety symptoms were stable from 30 months to 4–6 years.

**Conclusions:** Neural responsivity to unexpected stimuli in early infancy moderates developmental pathways from temperamental risk to childhood anxiety. Findings replicate and extend prior work to an earlier developmental window and a community sample, highlighting infancy as a critical period for identifying neurobiological markers of anxiety risk.

## Introduction

Anxiety disorders often emerge in childhood and exert long-lasting effects on development. Behavioral inhibition (BI), a temperament characterized by novelty-evoked avoidance (Henderson, Pine, & Fox, 2015), is a robust behavioral marker of risk for anxiety. BI is commonly preceded by negative reactivity (NR), a temperament characterized by distress in response to novelty (Kagan, 1997). Even so, only about 30% of infants with NR develop BI (Fox, Snidman, Haas, Degnan, & Kagan, 2015), and only about 40% of infants with BI ultimately develop anxiety disorders (Clauss & Blackford, 2012), highlighting the heterogeneity in the pathways linking early temperament to anxiety outcomes. Several factors moderating risk identified include executive skills (White, McDermott, Degnan, Henderson, & Fox, 2011), caregiving (Ryan & Ollendick, 2018), and neural indices of deviance or novelty (Filippi et al., 2020; Reeb-Sutherland et al., 2009; Xing et al., 2026). Most of these moderating factors exert their impact in childhood and adolescence. To date, only one study has shown that *infant* brain response to deviance influences risk trajectories both from NR to BI, and from BI to anxiety outcomes (Xing et al., 2026). Even so, this prior study was conducted in a cohort enriched for NR, leveraged EEG data at two distinct periods of infancy (9 and 36 months), and examined anxiety outcomes in adolescence. These features raise questions about both the generalizability of the observed effects to non-selected community samples, and the specificity of the effects to these developmental stages. The current study addresses these limitations by replicating the prior findings in an independent, community-sampled cohort and examining neural processes earlier in infancy in association with childhood anxiety outcomes.

Xing and colleagues (2026) measured EEG during a three-stimulus auditory oddball paradigm to assess how infants responded to unexpected sensory stimuli in the environment. During this paradigm, infants were presented with three types of auditory stimuli: repetitive and invariant “standard” stimuli (ie, pure tones); infrequent, unvarying “deviant” stimuli (e.g., tones differing in frequency from the standard) (Näätänen, 1995; Näätänen, 1990); or infrequent, qualitatively distinct “novel” stimuli (e.g., car horn) (Escera, Alho, Schröger, & Winkler, 2000; Escera, Alho, Winkler, & Näätänen, 1998). In infants, deviant stimuli after repeated standard stimuli typically elicit a positive ERP deflection around 150-250 ms after stimulus onset. This mismatch response (MMR), computed as the difference between responses to deviant and standard stimuli (deviant − standard), indexes a pre-attentive response to changes in stimulus properties (Garrido, Kilner, Stephan, & Friston, 2009). Novel stimuli typically elicit a positive ERP deflection around 300 ms after stimulus onset. The novelty P3, computed as the difference between novel and standard responses (novel − standard), indexes an orienting response to novelty (Kushnerenko, Van Den Bergh, & Winkler, 2013; Marshall, Reeb, & Fox, 2009; Reeb-Sutherland et al., 2009). Leveraging these event-related potentials (ERP) amplitudes, Xing and colleagues (2026) showed that infants with higher NR at 4 months and more negative MMR at 9 months were more likely to develop higher BI at 2-3 years; and those with higher BI at 2-3 years and more negative MMR at 3 years were more likely to develop greater anxiety symptoms as adolescents. Further, more positive MMR at 3 years were related to resilience, such that infants with higher BI and more positive MMR were more likely to show fewer anxiety symptoms in adolescence.

The current study pursues two directions to extend prior findings. First, it remains unclear whether specific developmental windows—such as 9 and 36 months—are uniquely critical for capturing meaningful variability in neural responses to deviance relevant to vulnerability to anxiety. This question is particularly relevant given that NR, as one of the earliest behavioral markers of temperamental risk for anxiety, emerges as early as 4 months of age (Kagan, 1997). Therefore, evaluating whether neural responsivity to deviant sensory changes at 4 months moderates early temperamental risk will inform whether infant risk prediction would benefit from comprehensive and combined behavioral and neural assessments at this early age. The current study leverages existing 4-month EEG data from the longitudinal Origins of Infant Temperament study (OIT) (Filippi et al., 2024) to extend effects revealed in prior work (i.e., Xing et al. (2026) to earlier developmental windows. Further, whereas prior work examined adolescent anxiety outcomes using gold-standard diagnostic measures (Xing et al., 2026), the current study tests whether similar effects are evident earlier in development in a younger cohort, using parent-reported measures of childhood symptoms of anxiety. Second, it remains unclear whether the original findings extend beyond the selectively sampled Temperament Over Time Study (TOTS) cohort (Xing et al., 2026), which intentionally overrepresented infants with extreme temperamental reactivity. Although this sampling strategy facilitated the identification of infants at elevated risk for BI and anxiety, it may also have amplified associations among temperament, neural measures, and later anxiety outcomes. Replication in a community sample would therefore clarify whether these effects generalize beyond infants selected based on early reactivity.

To address these gaps, the current study utilized data from the OIT study (Filippi et al., 2024), an independent cohort of community-sampled infants. The OIT study recruited 4-month-old infants who were observationally assessed for NR but not selected based on their reactivity scores. All infants were invited to participate in the same auditory oddball paradigm with concurrent high-density EEG recording at 4 months. Children were then followed prospectively, with BI observationally assessed at 14 months and anxiety outcomes measured via parent report on the Child Behavior Checklist (Achenbach & Rescorla, 2001) at 30 months, and again between 4 and 6 years of age. Structural equation modeling was used to evaluate the moderation effect of MMR on the pathways from NR to BI, and from BI to anxiety. In line with prior work, we hypothesized (1) that infants with higher NR and more negative MMR would show greater BI; and (2) that infants with higher BI and more negative MMR would exhibit greater childhood anxiety symptoms. Furthermore, following prior work (Xing et al., 2026), we tested whether more positive MMR might serve as a protective mechanism, such that infants with higher BI and more positive MMR would show fewer anxiety symptoms in childhood. Critically, this study directly replicates the analytic framework of Xing et al. (2026) but extends the study by using a community-sampled cohort and different developmental timepoints for EEG, BI, and anxiety assessments.

## Methods

### Participants

Participants were drawn from the OIT study, an ongoing longitudinal investigation examining the neurobiological foundations of early-life temperament via EEG and functional magnetic resonance imaging (fMRI) (Filippi et al., 2024). Exclusion criteria included preterm birth (< 37 weeks gestation), low birth weight (< 2500 g), significant birth complications, diagnosed neurological or developmental disorders, uncorrected visual or auditory impairments, MRI contraindications, and a history of neurological problems or head injury (see *Supporting Information* for details of recruitment). The final sample included *N*=147 infants (M_age at NR_ = 4.29 months; 65 females). Participants were predominantly White and Non-Hispanic (61.90%), and most mothers held advanced degrees (eg, master’s or PhD; 63.27%). Detailed demographic characteristics are presented in Table 1. Prior to data collection, parents provided written informed consent. Enrolled participants elected to complete EEG assessments, MRI assessments, or both. All study procedures were approved by the Institutional Review Board at University of Maryland, College Park.

**Table 1.** Sample demographics.

|  |  | **N (% of sample)** | **Mean (SD)** |
| --- | --- | --- | --- |
| *N* |  | 147 | - |
| Male |  | 82 (55%) | - |
| Age at Negative Reactivity Visit |  | - | 4.29 (0.56) |
| Age at EEG Visit |  | - | 5.18 (0.74) |
| Age at BI Visit |  | - | 14.4 (1.59) |
| Age at first CBCL assessment (in months) |  | - | 30.98 (3.4) |
| Age at second CBCL assessment (in years) |  | - | 4.81 (0.66) |
| Race/Ethnicity | African American/Hispanic | 1 (0.68%) |  |
|  | Caucasian/Hispanic | 12 (8.16%) |  |
|  | Multi-Racial/Hispanic | 3 (2.04%) |  |
|  | Asian/Hispanic | 1 (0.68%) |  |
|  | African American/Non-Hispanic | 5 (3.40%) |  |
|  | Caucasian/Non-Hispanic | 91 (61.90%) |  |
|  | Multi-Racial/Non-Hispanic | 20 (13.61%) |  |
|  | Asian/Non-Hispanic | 8 (5.44%) |  |
|  | Missing | 6 (4.08%) |  |
| Maternal Education | GED | 2 (1.36%) |  |
|  | High School Diploma | 1 (0.68%) |  |
|  | Some College | 8 (5.44%) |  |
|  | Two Year or Professional Degree | 4 (2.72%) |  |
|  | Four Year College Degree | 33 (22.45%) |  |
|  | Advanced Degree | 93 (63.27%) |  |
|  | Missing | 6 (4.08%) |  |

**Note.** CBCL refers to Child Behavior Checklist.

All *n* = 95 participants who chose to participate in EEG attempted an auditory oddball EEG task, with *n* = 5 participants excluded from subsequent analyses due to capping failure, excessive movement, or poor data quality (see EEG Data Processing for details). Participants who completed the auditory oddball task did not differ from those who did not in terms of sex, maternal education, or race/ethnicity (*p*s > .12). Little’s Missing Completely at Random (MCAR) test indicated that data were missing at random across time points (*p* = .89). Accordingly, all analyses utilized all available data, with missing data handled using full-information maximum likelihood (FIML) estimation (Enders & Bandalos, 2001).

### Negative Reactivity (NR)

NR was assessed at 4 months using the standardized Kagan reactivity assessment (Kagan & Snidman, 1991). Infants’ motor and affective responses to novel visual and auditory stimuli were observed across alternating visual and auditory blocks while sessions were video recorded for offline coding. Visual blocks consisted of three mobiles varying in number of hanging toys (1, 3, and 6), each presented three times for 20 seconds with 10-second intertrial intervals (nine trials per block). Auditory blocks included (1) eight short sentences (∼2 seconds each, 2-second intertrial interval) and (2) constant-vowel sounds (eg, ma, pa, ga) presented across three 10-second trials with 5-second intertrial intervals. Motor activity (e.g., leg kicks, arm waves), positive affect (e.g., smiling, laughter), and negative affect (e.g., fussing, crying) were rated on 7-point Likert scales. A second rater independently coded 95% of videos, demonstrating good inter-rater reliability (ICCs = 0.76–0.92; Shrout & Fleiss, 1979). Subscale scores were averaged across blocks and standardized. The NR composite was computed as the product of standardized negative affect and motor activity. Additional temperament measures and sensitivity analyses are reported in the *Supporting Information*.

### EEG Data Acquisition

At 4 months, EEG data were collected during a passive three-stimulus auditory oddball paradigm (see *Supporting Information* for full details). In short, the paradigm consisted of four blocks, each containing 400 auditory stimuli (∼3.5 minutes per block), with three stimulus categories: standard, deviant, and novel. Standard and deviant stimuli were pure tones composed of sine waves but differed in frequency (500 Hz vs 650 Hz). Tone frequencies were counterbalanced across blocks. Novel stimuli were complex, non-repeating sounds (eg, car horning, cow mooing) (Fabiani, Kazmerski, Cycowicz, & Friedman, 1996). Within each block, 80% of trials were standard tones, 10% were deviant tones, and 10% were novel sounds. Infants completed up to four blocks of the task, with a mean of 3.5 blocks completed (*SD* = 0.73).

### EEG Data Processing

EEG data were processed using the Maryland Analysis of Developmental EEG pipeline (Debnath et al., 2020; Leach et al., 2020) implemented in MATLAB (MathWorks, Natick, MA) using EEGLAB-based tools (Delorme & Makeig, 2004; Hunt, Lipsman, & Rosenberg, 2014). In brief, continuous data were filtered, and artifacts were identified and removed using automated and semi-automated procedures, including independent component analysis. Data were then segmented into epochs, baseline-corrected, and subjected to epoch-level artifact rejection. Remaining noisy channels were interpolated, and data were re-referenced to the average (see *Supporting Information* for full details).

The ERPs were computed separately for each stimulus condition. Based on inspection of the topographic distributions (see Figure 1) and MMR literature with infants (Themas, Lippus, Padrik, Kask, & Kreegipuu, 2023), we focused on frontolateral sites F3 and F4 in the current study (F3 cluster: E19, E20, E24, E27, E28, E29; F4 cluster: E4, E111, E117, E118, E123, E124). Participants with 20 or fewer usable trials in any condition were excluded (*n* = 3). At 4 months of age, the mean number of artifact-free trials contributing to ERP averages was 157.12 (*SD* = 63.87) for standard condition, 79.26 (*SD* = 32.24) for deviant condition, and 80.23 (*SD* = 32.03) for novel condition. The number of usable EEG data was not significantly associated with NR, BI, or anxiety outcomes (*p*s > .2). The MMR were calculated as the mean amplitude difference between deviant and standard conditions within the 100–300 ms post-stimulus time window (Deviant − Standard). The novelty P3 component was derived by subtracting responses to standard tones from responses to novel sounds in the 100–400 ms post-stimulus interval (Novel − Standard). These difference scores isolate neural sensitivity to auditory deviance and novelty, respectively. Grand-average ERPs averaged across F3 and F4 clusters by stimulus type at 4 months, along with corresponding scalp topographies, are presented in Figure 1.

**Figure 1.**
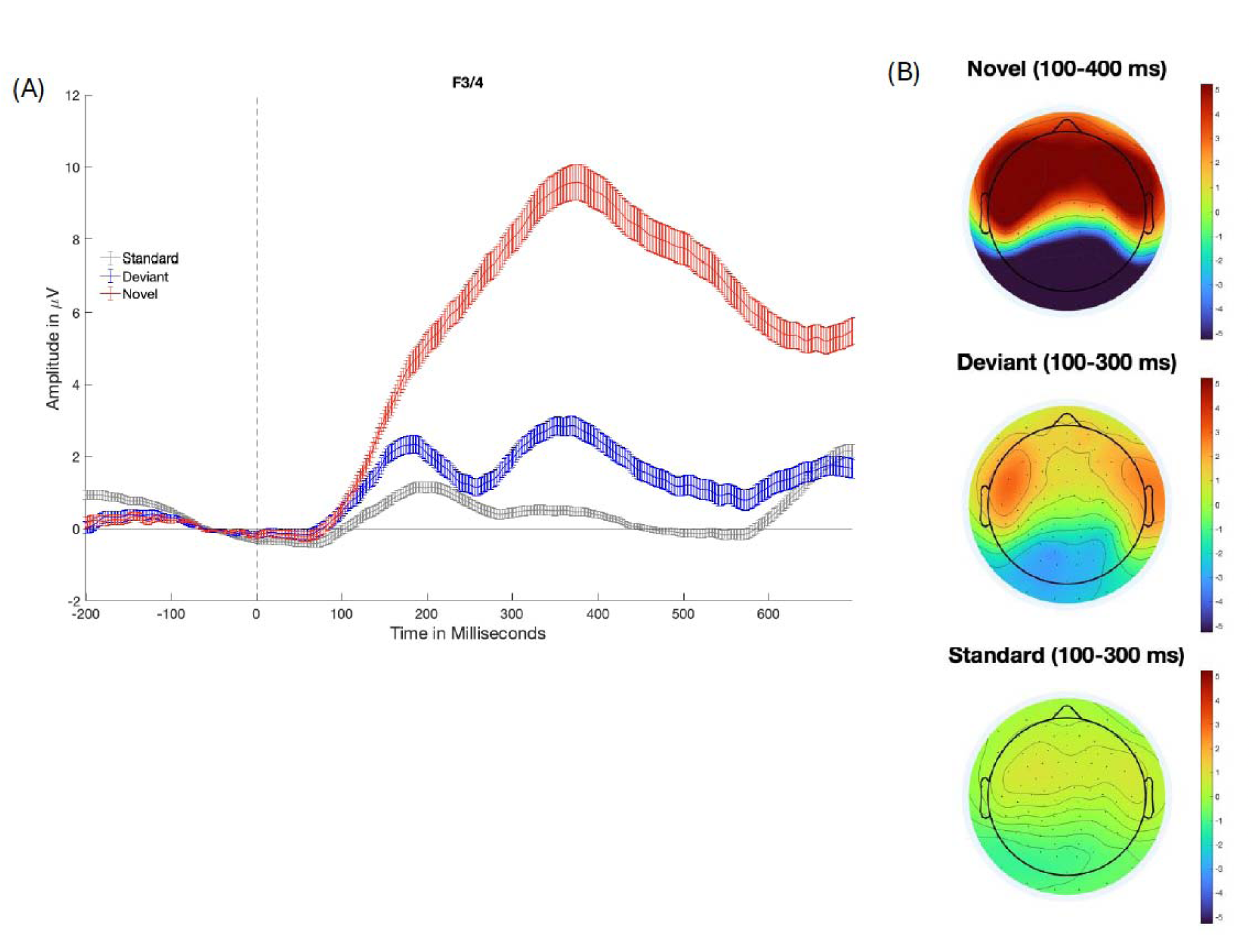
(A) ERP waveforms for standard, deviant and novel conditions averaged across F3 and F4 clusters (*n*= 90); (B) The scalp topographic maps of neural responses in each condition. **Behavioral Inhibition (BI)**

At 14 months, BI was assessed using a standardized observational paradigm (Kagan et al., 1984). Infants’ responses to unfamiliar stimuli were observed across three structured episodes involving a stranger, a motorized robot, and an inflatable tunnel. Sessions were videotaped for offline coding of latency to approach or vocalize and proximity to the mother. Indicators were standardized and averaged across episodes to derive a composite BI score. Among infants with BI data (*n* = 64), 36% of videos were double-coded, demonstrating high interrater reliability (mean ICC = 0.95). Additional parent-report measures and sensitivity analyses are reported in the *Supporting Information*.

### Child Behavior Checklist

The Child Behavior Checklist (CBCL) (Achenbach & Rescorla, 2001), a standardized parent-report questionnaire of child behavior, was administered at 30 months and 4–6 years. Parents rated 99 items describing child behaviors on a 3-point scale (0 = *Not True*, 1 = *Somewhat or Sometimes True*, 2 = *Very True or Often True*). Anxiety symptoms were indexed using the DSM-oriented *Anxiety Problems* subscale, which served as the primary outcome. *Attention Problems* and *Externalizing Problems* subscales were included as covariates in sensitivity analyses to assess specificity. To address the influence of the equal item contribution (tau-equivalence) assumption and sample size on estimates of internal consistency, McDonald’s omega was calculated for CBCL scores (Dunn, Baguley, & Brunsden, 2014; McDonald, 1999; Orcan, 2023). Internal consistency was good to excellent at 30 months (ω = 0.90 for *Anxiety Problems*, ω = 0.88 for *Attention Problems*, ω = 0.84 for *Externalizing Problems*) and acceptable to good at 4–6 years (ω = 0.79, 0.70, and 0.76, respectively).

### Analytic Approach

Correlations among all variables of interest were first examined (see Table 2). To test the hypotheses, we leveraged all available data and conducted structural equation modeling (SEM) using the “lavaan” package (Rosseel, 2018) in R (version 4.4.1), with missing data handled with FIML (Enders & Bandalos, 2001). Model fit was evaluated with multiple indices, including the Chi-square (χ²) test, the root-mean-square error of approximation (RMSEA), the comparative fit index (CFI), and the standardized root mean square residual (SRMR). All variables were standardized (z-scored) prior to SEM. The focal SEM model examined the interaction between MMR and NR at 4 months in predicting BI at 14 months, the interaction between MMR at 4 months and BI at 14 months in predicting anxiety outcomes at 30 months, and associations between anxiety outcomes across 30 months and 4-6 years. Only statistically significant paths were retained in the final model to optimize model fit. Standardized parameter estimates, standard errors, and statistical significance were reported for all included paths (See Figure 2).

**Figure 2.**
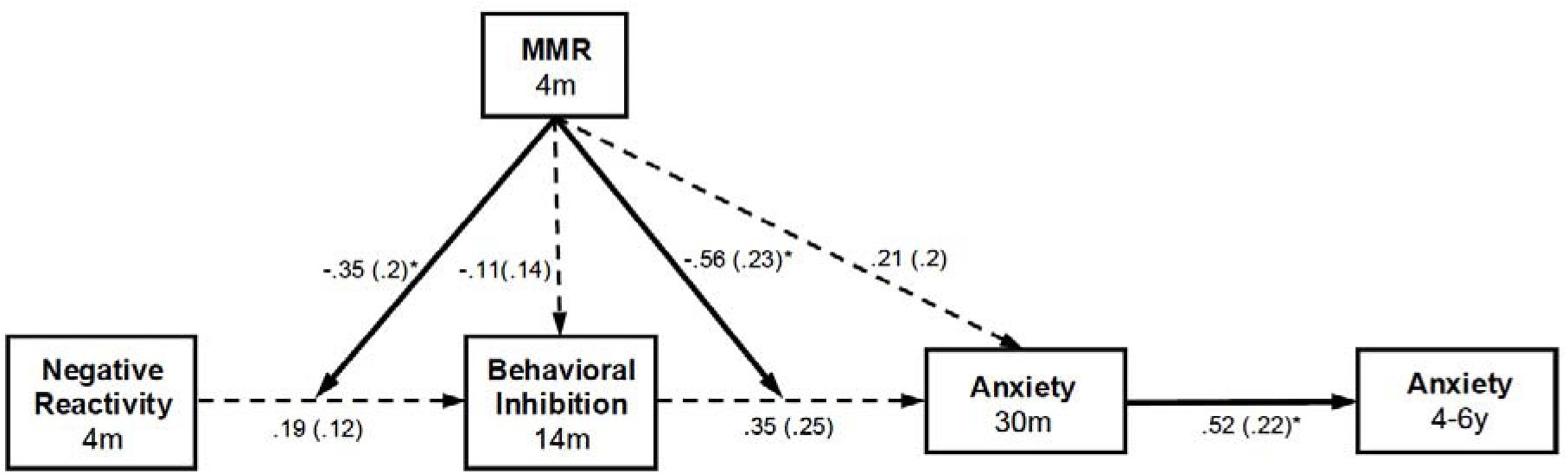
Focal structural model tested. Standardized parameter estimates are shown with standard error in parentheses. Statistically significant (*p* < .05) paths are indicated with solid lines and asterisks while non-significant paths are indicated with dashed lines. \**p* < .05.

**Table 2.** Means, standard deviations, and correlations with confidence intervals.

| Variable | *n* | *M* | *SD* | 1 | 2 | 3 | 4 | 5 |
| --- | --- | --- | --- | --- | --- | --- | --- | --- |
| 1. 4m NR | 130 | 0.32 | 1.22 |  |  |  |  |  |
| 2. 4m P3 | 90 | 4.9 | 2.91 | 0.15<br>[-0.06,<br>0.35] |  |  |  |  |
| 3. 4m<br>MMR | 90 | 0.69 | 2.16 | -0.11<br>[-0.31,<br>0.11] | 0.31**<br>[0.11,<br>0.49] |  |  |  |
| 4. 14m BI | 64 | -0.02 | 0.57 | 0.20<br>[-0.41,<br>0.43] | 0.07<br>[-0.22,<br>0.35] | -0.12<br>[-0.39,<br>0.17] |  |  |
| 5. 30m<br>Anxiety | 23 | 2.9 | 1.98 | -0.15<br>[-0.53,<br>0.29] | -0.12<br>[-0.54,<br>0.34] | 0.25<br>[-0.22,<br>0.62] | 0.41<br>[-0.15,<br>0.77] |  |
| 6. 4-6y<br>Anxiety | 74 | 2.8 | 2.61 | -0.17<br>[-0.39,<br>0.06] | -0.01<br>[-0.31,<br>0.28] | 0.18<br>[-0.12,<br>0.45] | 0.13<br>[-0.18,<br>0.41] | 0.41<br>[-0.04,<br>0.72] |

*Note.* NR represents negative reactivity, BI represents behavioral inhibition, P3 refers to the novelty P3 response, and MMR refers to the mismatch response. *M* and *SD* are used to represent mean and standard deviation, respectively. Values in square brackets indicate the 95% confidence interval for each correlation. The confidence interval is a plausible range of population correlations that could have caused the sample correlation. \* indicates $p < .05$ .

For significant moderation effects, traditional Johnson-Neyman procedures could not be directly applied due to sample size limitations and missing data considerations. Accordingly, regions of significance were derived using model-implied parameter estimates from the SEM, following Preacher et al. (2006). Specifically, conditional effects of the predictor on the outcome were computed across values of the moderator (MMR), and their standard errors were estimated using the variance-covariance matrix of the model parameters. This approach, analogous to the Johnson-Neyman technique, was used to identify the range of MMR values at which (a) NR was significantly associated with BI; and (b) BI was significantly associated with anxiety outcomes.

We additionally confirmed that NR was not directly associated with anxiety outcomes across 30 months and 4-6 years (*p*s > .14), nor did NR interact with MMR at 4 months to predict anxiety across time (*p*s > .86). Thus, these non-significant paths were excluded from the final SEM. Further, although prior work (Xing et al., 2026) demonstrated a significant mediation effect of P3 linking NR and anxiety outcome, preliminary correlational analyses indicated non-significant associations (1) between NR and 4-month novelty P3 (*p* = .16), and (2) between 4-month novelty P3 and anxiety across time points (*p*s > .6), precluding the replication of these mediation models. Additionally, the lack of significant associations between MMR amplitudes and NR, BI, and anxiety outcomes across time (*p*s > .23) precluded testing mediation models in which MMR serves as a mediator between NR and BI, or between BI and anxiety outcomes.

To evaluate the specificity of the MMR effects, several sensitivity analyses were conducted. First, the focal SEM model was replicated using novelty P3 as the moderators. Second, the model was replicated with *Attention Problems* and *Externalizing Problems* subscales from CBCL as covariates in predicting anxiety outcomes, which provided evidence for the specificity of the risk trajectories to anxiety. In the *Supporting Information*, we additionally present the following sensitivity analyses on: (1) robustness of findings to different operationalizations of temperament, assessed by replicating the focal SEM model using alternative measures of NR and BI; (2) exploratory analyses examining whether the rate of changes in neural responses to deviance across time could account for the focal findings, following Xing et al. (2026); and (3) sensitivity analyses using MMRs extracted at Fz.

## Results

The MMR amplitudes at 4 months significantly moderated the association between NR at 4 months and BI at 14 months (β = -.35, *p* = .04). The amplitude of the MMR at 4 months also significantly moderated the association between BI at 14 months and anxiety outcome at 30 months (β = -.46, *p* = .04). The anxiety outcomes across 30 months and 4-6 years were also significantly associated (β = .49, *p* =.03). The SEM model revealed good model fit (χ^2^ (8) = 4.08, *p* = .85; CFI = 1; RMSEA = 0; SRMR = .06) (Hu & Bentler, 1999) (See Figure 2). Post-hoc probing analyses using model-implied conditional effects revealed that higher NR at 4 months related to greater BI at 14 months, but only among infants with more negative MMR at 4 months (*p*< .05, z-score range [-3.61, -0.09]). Infants with more positive MMR at 4 months (z-score range [-0.09, 2.69]) showed no significant associations between NR and BI (See Figure 3(A)).

**Figure 3.**
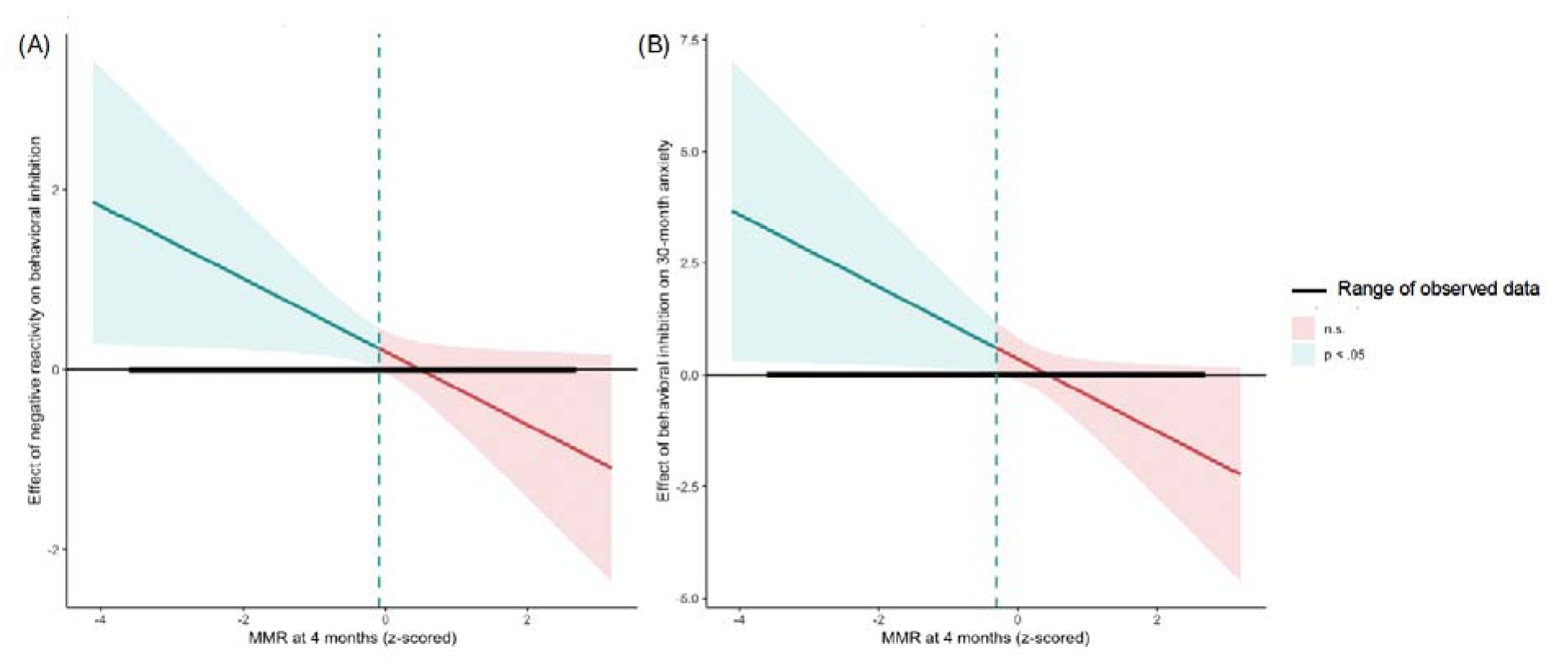
Regions of significance plots for the focal model. The blue and red shaded areas represent the levels of the moderator variable, mismatch response (MMR) when the associations are statistically significant (*p* < .05) and non-significant (*p* > .05), respectively. Figures illustrate that infants with more negative MMR at 4 months show a positive association (A) between negative reactivity and behavioral inhibition, and (B) between behavioral inhibition and anxiety, whereas no significant associations emerged at more positive MMR values.

Similarly, higher BI related to greater anxiety at 30 months, but only among infants with more negative MMR at 4 months (*p*< .05, z-score range [-3.61, -0.55]). Infants with more positive MMR at 4 months (z-score range [-0.55, 2.69]) showed no significant associations between BI and anxiety (See Figure 3(B)).

### Sensitivity analyses

Parallel analyses using novelty P3 response as the moderators yielded non-significant moderation effects (*p*s > .72). The moderating effect of MMRs on the association between BI and anxiety outcome at 30 months remained significant, when controlling for attention problems (β = .52, *p* =.03) or externalizing problems (β = .61, *p* <.001), despite a decrease in model fit. Specifically, model fit indices were χ^2^ (10)= 6.73, *p*= .75; CFI= 1; RMSEA= 0; SRMR = .07 when controlling for attention problems; and were χ^2^ (10)= 9.61, *p*= .48; CFI= 1; RMSEA= 0; SRMR = .11 when controlling for externalizing problems.

## Discussion

The present study leveraged existing longitudinal data to investigate whether neural responsivity to unexpected sensory stimuli in early infancy moderated developmental pathways from temperamental risk to childhood anxiety. Findings indicated that a more negative MMR at 4 months, reflecting heightened neural sensitivity to sensory deviance, was related to an likelihood that infants with high NR at 4 months would maintain temperamental risk and develop high BI at 14 months. Similarly, a more negative MMR was related to an likelihood that infants with high BI at 14 months would exhibit elevated anxiety symptoms in childhood. These results extend prior work implicating infant pre-attentive change detection in anxiety risk (Xing et al., 2026) and demonstrates that the MMR measured as early as 4 months influence pathways to childhood anxiety. The study highlights the importance of integrating behavioral markers with neural indices in early infancy to improve the identification of emerging anxiety-related vulnerability.

Our findings are robust and consistent with prior work (Xing et al., 2026). Despite a substantial generational gap between cohorts (TOTS recruited in the late 1980s to 1990s, and OIT recruited in the late 2010s), consistent effects emerged. Replication in a community sample (i.e., OIT) demonstrates generalizability and extends prior work to earlier EEG and outcome timepoints. The consistency in MMR moderating risk trajectories across 4 months, 9 months, and 36 months could suggest that alterations in pre-attentive processing as a relatively stable indicator of risk across infancy. However, longitudinal studies are needed to confirm stability.

Together, our data highlight that pre-attentive processes shape anxiety trajectories earlier than previously recognized (Reeb-Sutherland et al., 2009; Xing et al., 2026), underscoring the value of early infancy as a window for early identification of risk for anxiety.

We did not observe the resilience effect reported in prior work (Xing et al., 2026), in which more positive MMR indexed a protective mechanism linking higher BI to lower anxiety symptoms. This difference may be attributable to a narrower distribution of anxiety symptoms in childhood (z score range: -1.06 to 2.49) compared to that previously reported in adolescence (z score range: -1.43 to 4.75) in TOTS (Xing et al., 2026), which may reflect less severe or less differentiated anxiety symptoms in early childhood. The smaller range of anxiety symptoms, paired with the smaller sample size, may have reduced the statistical power.

The directionality of the MMR moderation effects was consistent with our hypothesis. Building from work in older populations showing that larger (i.e., more negative) mismatch negativity (MMN) may reflect larger prediction error (Garrido et al., 2009; Wacongne, Changeux, & Dehaene, 2012), our data may suggest that at-risk infants show a larger prediction error or are more sensitive to the discrepancy between the expected vs actual sensory input.

Indeed, those behaviorally inhibited infants who developed high anxiety showed increasingly negative MMR over the course of the experiment, suggesting larger increase in prediction errors. In contrast, at-risk infants whose MMR became increasingly positive with repeated presentations of deviance, suggesting larger decrease in prediction errors, showed reduced childhood anxiety—suggestive of a resilience pathway. These findings may implicate the heightened sensory processing of deviance as an early-life precursor to heightened error-related processing associated with anxiety (Filippi et al., 2020; Meyer, 2017), but additional work is needed to evaluate this possibility. This effect was specific to the BI to anxiety pathway and not observed for pathways from NR to BI (see Xing et al., 2026, which observed the opposite pattern).

Differences in sample characteristics, developmental timing, and task parameters may have all contributed to these discrepancies, underscoring the need for further investigation. Together, these findings underscore the importance of replication across samples and developmental stages evaluate the specificity of these pathways.

Across the TOTS and OIT cohorts, the moderation effects were specific to the MMR and were not observed for the novelty P3 response. In this replication, we did not find that the 4-month novelty P3 mediated the pathway from NR to anxiety (a finding previously reported on the TOTS cohort (Xing et al., 2026)) nor did the novelty P3 moderate pathways from BI to anxiety (as has been shown in adolescence (Reeb-Sutherland et al., 2009)). The absence of a P3 mediation effect in the current study may reflect the still-limited maturation and stability of P3 in early infancy, reducing its ability to reliably mediate or moderate risk pathways at 4 months.

These discrepancies highlight the need for longitudinal studies to clarify developmental changes in both MMR and P3.

The present study has several notable strengths. First, its longitudinal design beginning at 4 months, providing prospective evidence of developmental pathways from early neural and temperamental risk markers to later anxiety outcomes. Second, the use of observational, gold-standard assessments of NR and BI during infancy, strengthens the validity of the findings. Third, the specificity of effects to observed measures of NR and BI further underscores their utility in capturing individual differences in early temperament relative to parent-report measures (e.g., IBQ, TBAQ) or less comprehensive operationalizations (e.g., negative affect alone). Fourth, anxiety outcomes were assessed in early childhood (30 months and 4–6 years), aligning with emerging large-scale efforts (e.g., HBCD) and facilitating future replication in more diverse samples. Finally, relative to prior work (Xing et al., 2026), which relied on EEG data collected using a low-density system (14 channels), the present study employed high-density EEG (128 channels) along with an expanded task design featuring a greater number of blocks. This methodological choice provide finer spatial sampling, improves the signal-to-noise ratio (Michel, 2019), and enhances measurement reliability.

Several limitations should also be considered. Firstly, although MMR effects remained significant after covarying for attention and externalizing problems, specificity to anxiety is uncertain. Some high correlations (r>.5) among CBCL subscales (see Table S1) limit our ability to differentiate symptom domains and confidence in outcome specificity. This is further constrained by reliance on non-diagnostic parent-report measures and limited availability of gold-standard clinical assessments in early childhood. This limitation highlights the need for detailed characterization of early emerging psychopathology in longitudinal studies. Secondly, moderate level of data missingness, along with restricted variability and small sample size in anxiety symptoms at 30 months, may have reduced statistical power. Thirdly, the sample also lacked socioeconomic and racial diversity, limiting generalizability. Finally, the developmental trajectories of MMR and novelty P3 across infancy and early childhood remain incompletely understood, and how these processes dynamically shape risk pathways over time warrants further investigation. Addressing these gaps will be critical for clarifying the developmental specificity, stability, and robustness of MMR as a neural marker of anxiety risk.

## Conclusion

In summary, the present study provides evidence that neural responsivity to unexpected sensory stimuli (as indexed by the MMR) influences developmental pathways from early temperamental risk to later childhood anxiety. This work extends prior work and highlights early infancy as a key window for identifying neurobiological markers of risk. Together, this work advances our understanding of early-emerging neural mechanisms underlying anxiety risk. This work provides initial evidence that the MMR could serve as a marker of risk or as a neural target for early intervention.

## Ethics Statement

Prior to data collection, parents provided written informed consent. All study procedures were approved by the Institutional Review Board at University of Maryland, College Park (IRBnet ID: 817133-98; approval effective 01/13/2026).

## Key points and relevance

### What’s known?

- Temperamental risk factors, including negative reactivity (NR) and behavioral inhibition (BI), are well-established predictors of anxiety, but not all at-risk infants develop later anxiety symptoms.

### What’s new?

- This study demonstrates that neural responsivity to unexpected sensory stimuli, indexed by the mismatch response (MMR) at 4 months, moderates developmental pathways from NR to BI and from BI to childhood anxiety.
- Findings replicate prior work in an independent community sample and extend effects to an earlier developmental window and childhood anxiety outcomes.

### What’s relevant?

- Results highlight infancy as a sensitive period for integrating behavioral and neural markers to improve early identification of anxiety-related vulnerability.
- These findings inform evaluating infant neural markers as potential targets for early risk identification and intervention approaches for anxiety risk.

## Data Availability

Jiayin Xing had full access to all the data and takes responsibility for the integrity of the data and the accuracy of the data analysis. The data supporting the findings of this study, as well as analysis code and study materials, are available from the corresponding author upon reasonable request.

## Acknowledgement

This research was supported by the National Institutes of Health (NIH) under grants R21MH122976 (PI: Fox) and R00MH125878 (PI: Filippi). Dr. Daniel Pine’s contributions were supported in part by the National Institute of Mental Health Intramural Research Program (Project ZIA-MH002782). The contributions of NIH author(s) are considered works of the United States Government. The findings and conclusions in this paper are those of the authors and do not necessarily reflect the views of the NIH or the U.S. Department of Health and Human Services. We thank the children and families who participated in the study.

## Declaration of interests

The authors report no potential conflicts of interest.

## Abbreviations

BI: behavioral inhibition
EEG: electroencephalography
MMR: mismatch response
NR: negative reactivity.

## Notes

### Competing Interest Statement

The authors have declared no competing interest.

### Author Declarations

All study procedures were approved by the Institutional Review Board at University of Maryland, College Park (IRBnet ID: 817133-98; approval effective 01/13/2026).

