## Supplemental Material for "Neural Responsivity to Unexpected Stimuli Moderates a Developmental Pathway to Anxiety: A Community-Sample Replication in Early Infancy and Childhood"

**Recruitment**

Between July 2018 and the present, two cohorts of typically developing infants were enrolled: a resting-state fMRI (rs-fMRI) cohort (*n* = 111), in which imaging primarily consisted of rs-fMRI; and a task-based fMRI cohort (*n* = 36), in which the study team piloted a novel three-stimulus auditory oddball task in the scanner. Importantly, inclusion and exclusion criteria were identical across cohorts, and all observational assessments, parent-report measures, and EEG data were collected using identical procedures and equipment. While all infants in the rsfMRI cohort (*n* = 111) and most infants in the task-based fMRI cohort (*n* = 28) were recruited between 4 and 7 months of age, a small subset of infants in the task-based fMRI cohort (*n* = 8) were recruited as newborns. Even so, the two cohorts did not differ in age at the negative reactivity or EEG assessments (*p*s > .08), supporting the integration of the samples in subsequent analyses. Recruitment methods included community-based outreach such as social media advertisements, outreach events, mailed invitations, and local parenting groups.

**EEG Data Acquisition**

At 4 months, EEG was recorded using a 128-channel HydroCel Geodesic Sensor Net and digitized at a sampling rate of 500 Hz using EGI Net Station software (Version 4; Electrical Geodesics, Inc., Eugene, OR). Prior to data collection, electrode impedances were assessed and verified to be below 100 kΩ across all channels. EEG data were collected during a passive three-stimulus auditory oddball paradigm (see *Supporting Information* for full details). In short, the paradigm consisted of four blocks, each containing 400 auditory stimuli (~3.5 minutes per block), with three stimulus categories: standard, deviant, and novel. Standard and deviant stimuli were pure tones composed of sine waves but differed in frequency (500 Hz vs 650 Hz). Tone frequencies were counterbalanced across blocks. For example, in Block 1, the 500 Hz tone served as the standard and the 650 Hz tone as the deviant. In Block 2, this assignment was reversed, with the 650 Hz tone as the standard and the 500 Hz tone as the deviant. This alternating pattern continued across the remaining blocks. Novel stimuli were complex, non-repeating sounds (eg, car horning, cow mooing) (Fabiani, Kazmerski, Cycowicz, & Friedman, 1996). Within each block, 80% of trials were standard tones, 10% were deviant tones, and 10% were novel sounds. Auditory stimuli (200 ms duration; 300 ms interstimulus interval) were delivered through free-field speakers at 75 dB peak sound pressure level while infants viewed a silent video. Infants completed up to four blocks of the task, with a mean of 3.5 blocks completed (*SD* = 0.73). EEG data acquisition and preprocessing procedures reported here were largely consistent with prior reports on this cohort, except for the processing related to source localization (Kanel et al., 2025). Additionally, this prior study included only participants with both EEG and MRI data from the rs-fMRI cohort (*n* = 25) (Kanel et al., 2025), whereas the present study used all usable EEG data from the full OIT sample.

**EEG Data Processing: MADE pipeline**

EEG data were processed using the Maryland Analysis of Developmental EEG pipeline (Debnath et al., 2020; Leach et al., 2020) implemented in MATLAB (MathWorks, Natick, MA) using EEGLAB-based tools (Delorme & Makeig, 2004; Hunt, Lipsman, & Rosenberg, 2014). Following prior infant EEG research (Colomer et al., 2023; Debnath, Salo, Buzzell, Yoo, & Fox, 2019; Mariscal et al., 2021), electrodes located along the periphery of the sensor net were excluded due to their heightened susceptibility to ocular, facial, and head movement artifacts, leaving a total of 104 channels retained for subsequent analyses.

Continuous EEG data were first high-pass filtered at 0.3 Hz and low-pass filtered at 50 Hz. Channels exhibiting excessive noise were automatically detected and removed using the FASTER plug-in in EEGLAB (Nolan, Whelan, & Reilly, 2010). The independent component analysis (ICA) was further applied to address ocular and muscle-related artifacts. ICA was performed on a duplicate version of the dataset that had been high-pass filtered at 1 Hz and segmented into 1-second epochs to optimize ICA decomposition. Additionally, noisy data segments were identified and rejected using a combined voltage threshold (±1000 μV) and spectral threshold (−100 to +30 dB) within the 20–40 Hz frequency range to target EMG-like activity. Channels with more than 20% of epochs rejected were removed. ICA weights were then applied (Debener, Thorne, Schneider, & Viola, 2010), and artifactual components were identified and removed using a semi-automated approach that combined the Adjusted-ADJUST algorithm (Leach et al., 2020) with manual inspection of component properties.

Following ICA-based artifact correction, the data were segmented into epochs of 1500 ms, spanning from −500 ms to 1000 ms relative to stimulus onset. Baseline correction was applied using the mean voltage in the −100 ms to 0 ms pre-stimulus interval. Following the baseline correction, epoch-level artifact rejection was conducted excluding epochs with voltages at frontal electrodes exceeding ±150 μV (which reflect ocular contamination). For epochs in which non-frontal channels exceeded ±150 μV, those channels were interpolated at the single-epoch level. Epochs containing more than 10% interpolated channels were rejected. Participants for whom all epochs were rejected were excluded from further ERP analyses. After these steps, remaining missing channels were interpolated using spherical spline interpolation, and the data were re-referenced to the average of all electrodes.

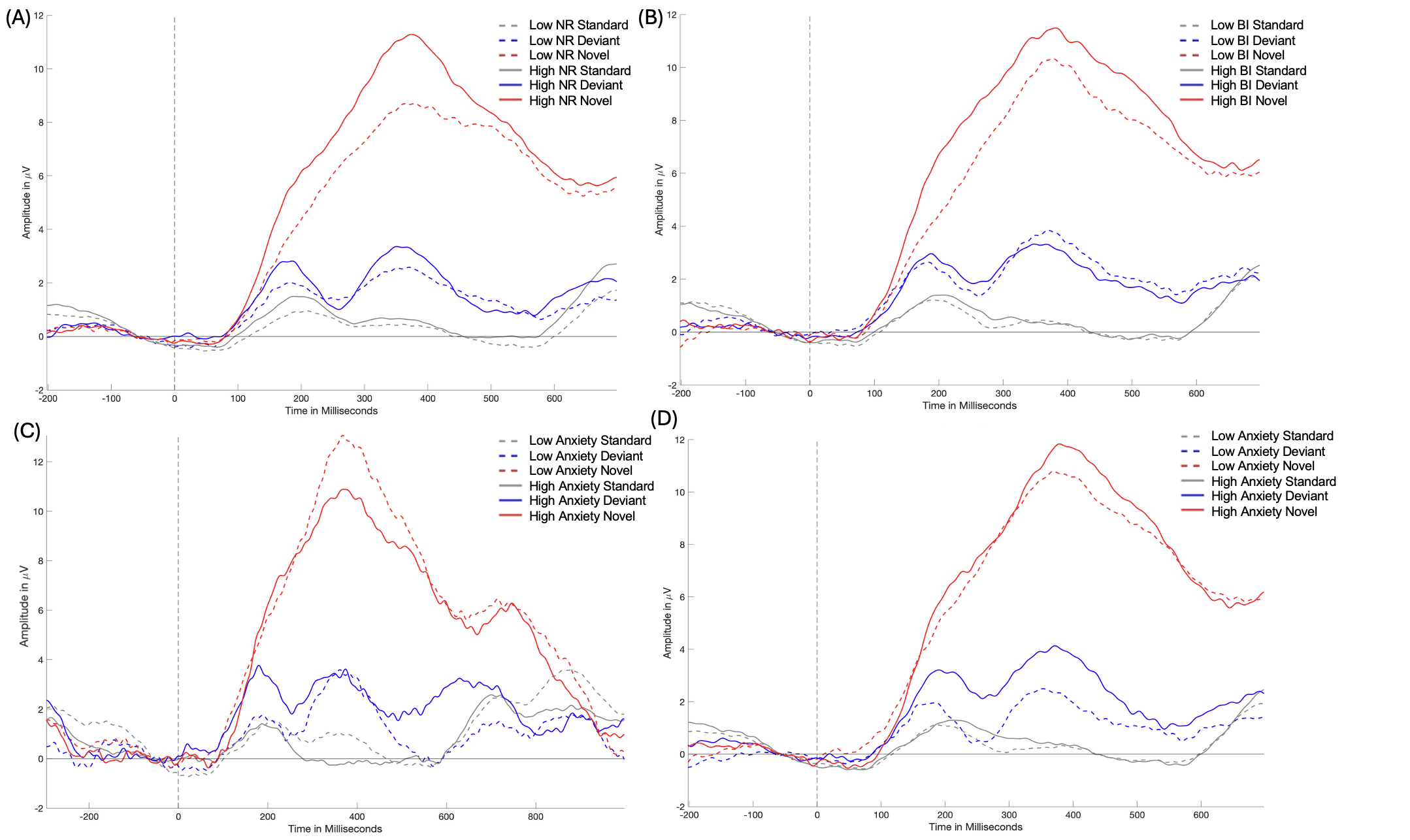

**Figure S1.** The grand mean ERPs to standard, deviant, and novel sounds at F3/F4 electrode clusters at 4 months, split by median negative reactivity (A), behavioral inhibition (B), 30-month anxiety (C), and 4-6-year anxiety (D).

*Note*. This figure is presented for illustrative purpose only. All focal analyses were conducted using continuous measures.

**Exploratory analysis: evaluating neural habituation to deviance at the block-level.**

According to the predictive coding frameworks established in adulthood (Marais & Roche-Labarbe, 2025), larger negative responses to deviant stimuli (MMN) are thought to reflect stronger prediction error signals when sensory input violates expected regularities, or a failure to attenuate responses and update expectations in response to incoming sensory changes. To test whether similar mechanisms operate in infancy, the rate of changes in neural responses to repeated deviance, indexing the modulation of prediction error in response to repeated deviant sensory stimuli across time, was examined in Xing et al. (2026) in association with MMR amplitudes and risk trajectories from temperamental risk to anxiety. Changes in neural responses to repeated deviant sounds during the auditory oddball paradigm has been documented in prior work in later development (McGee et al., 2001; Merchie & Gomot, 2023). Xing et al. (2026) showed that a more negative slope in neural responses to deviance, representing increasing or heighted prediction error across time, was associated with the maintenance of temperamental risk from NR to BI, driving the moderation effect of more negative MMRs at 9 months on risk trajectory. Further, a more positive slope, representing a greater attenuation in prediction error across time at 9 months, was associated with a resilience pathway from high BI to low anxiety. The current study aimed to expand on these results, to examine the rate of changes in neural responses to deviance at 4 months across time, and the associations with MMR amplitudes and the risk trajectory to anxiety.

The rates of change in neural responses to repetitive stimuli across 4 blocks were extracted by condition, using a series of mixed-effects linear regression models. The ‘lme4’ package (Bates et al., 2015) in R (version 4.1.0) was utilized, with block number predicting average amplitudes of neural responses, and subject-level random effects included to account for individual variability in neural response trajectories. Figure S2 presents the spaghetti plots of changes in neural responses across 4 blocks for the three conditions, shown separately for the two time windows from which MMR and P3 responses were extracted (100–300 ms and 100–400 ms, respectively). Red lines represent the fitted linear models.

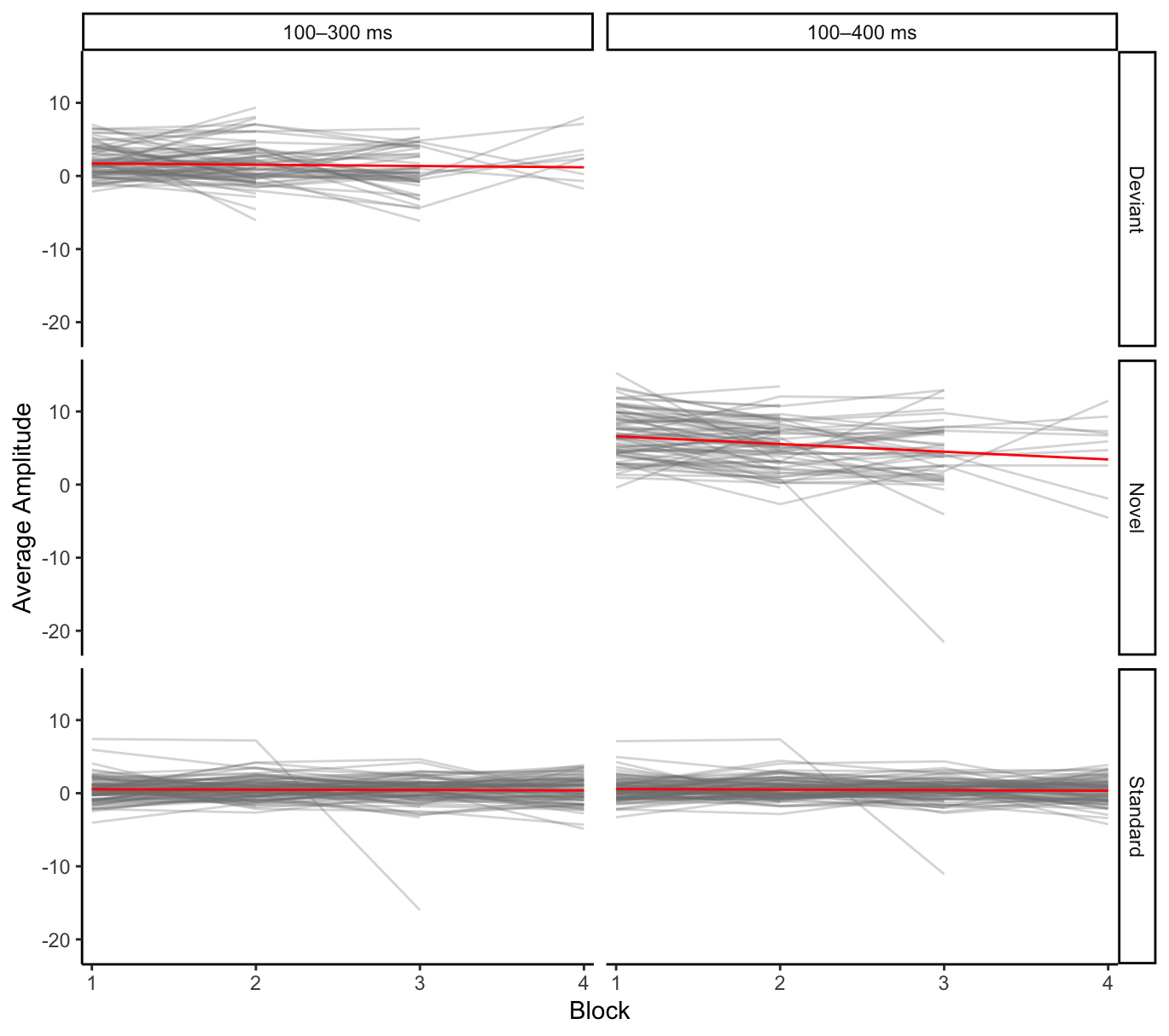

**Figure S2.** Change in neural responses across blocks, by condition and time window.

A significant effect of block/time on the amplitude of the neural responses was found for the novel condition (*Estimate* = -1.05, *p <* .001), and the standard condition (100-400 ms time window: *Estimate* = .06, *p =* .03; 100-300 ms time window: *Estimate* = .09, *p =* .003), but not for the deviant condition (*Estimate* = -.18, *p =* .4). Despite nonsignificant main effect of time/block, we examined individual differences in the extracted slope estimates of responses to deviant stimuli (i.e., rate of change), in association with MMR amplitudes, with the slope of neural responses to standard stimuli covaried as baseline. Consistent with our hypothesis and prior literature (Xing et al., 2026), a more negative slope (which we interpret in line with adult work as a larger increase in prediction error over time), was significantly correlated with more negative MMRs (*r*(87) = .66, *p <* .001) (see Figure S3).

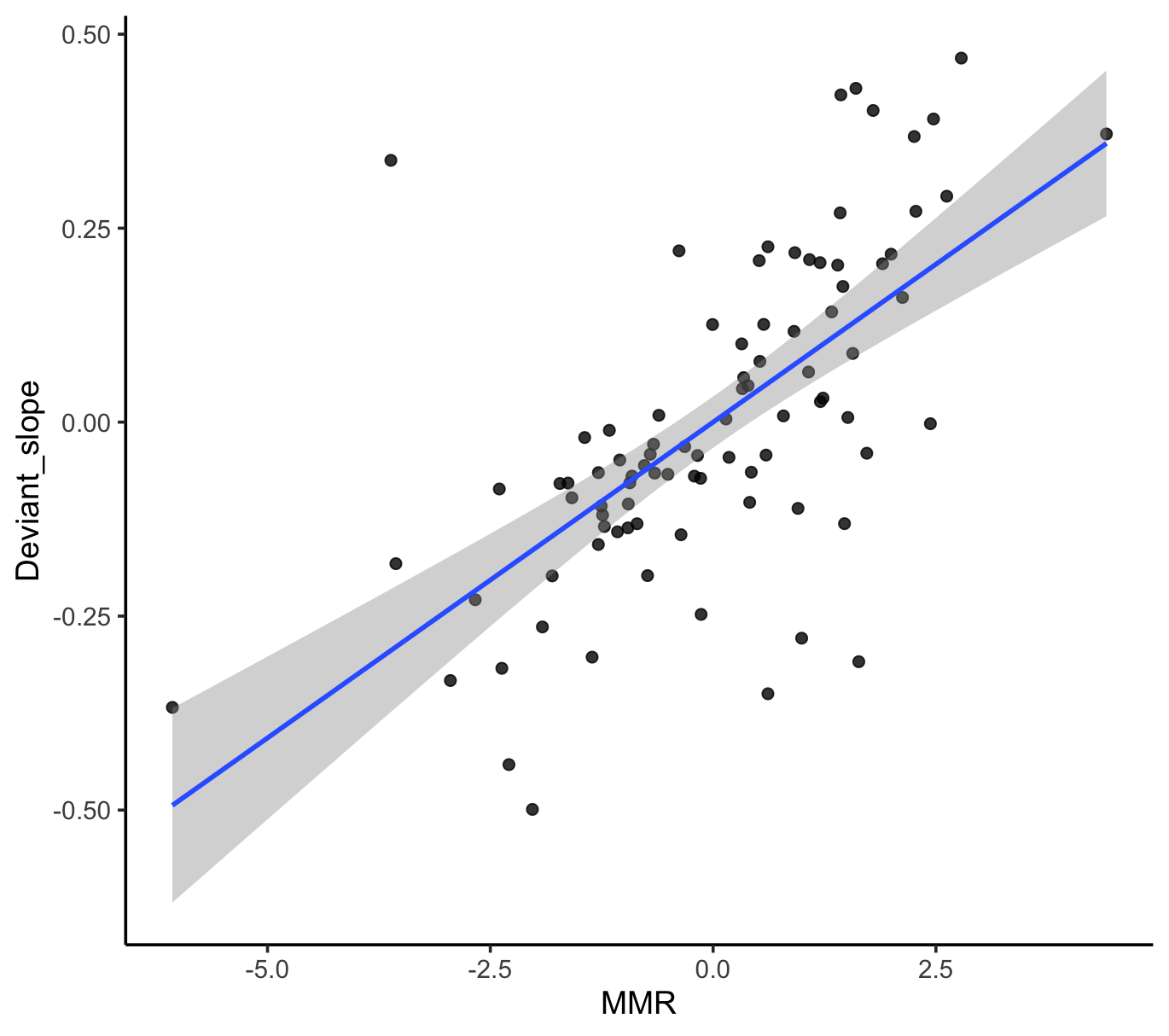

**Figure S3.** Correlations between the slope of neural responses to deviance and MMR amplitude, with the slope of neural responses to standard stimuli covaried.

We further examined whether the individual differences in rate of changes in neural responses to deviance may have driven the moderation effects of MMRs on risk trajectories to anxiety. To do so, the focal SEM model was replicated, by replacing MMR amplitudes with slopes indexing rate of changes in neural responses to deviant sounds, while also controlling for the slopes of neural responses to standard stimuli. Results indicated the slope of neural responses to deviance did not significantly moderate the association between NR at 4 months and BI at 14 months (*β* = -.07, *p* = .67), but significantly moderated the association between BI at 14 months and anxiety outcome at 30 months (*β* = -.57, *p* = .007), with a moderate model fit: χ^2^(9) = 10.47, *p* = .31; CFI = .81; RMSEA =.04; SRMR = .09. Post-hoc probing analyses using model-implied conditional effects revealed that higher BI at 14 months related to greater anxiety at 30 months, but only among infants with more negative slopes at 4 months (*p <* .05, z-score range [-2.12, -0.36]). Further, more positive slopes at 4 months (*p <* .05, z-score range [1.98, 2.44]) were associated with a resilience pathway from high BI at 14 months to lower anxiety at 30 months. No association was found between BI and anxiety for infants with medium level of slopes (*p >* .05, z-score range [-0.36, 1.98]) (See Figure S4).

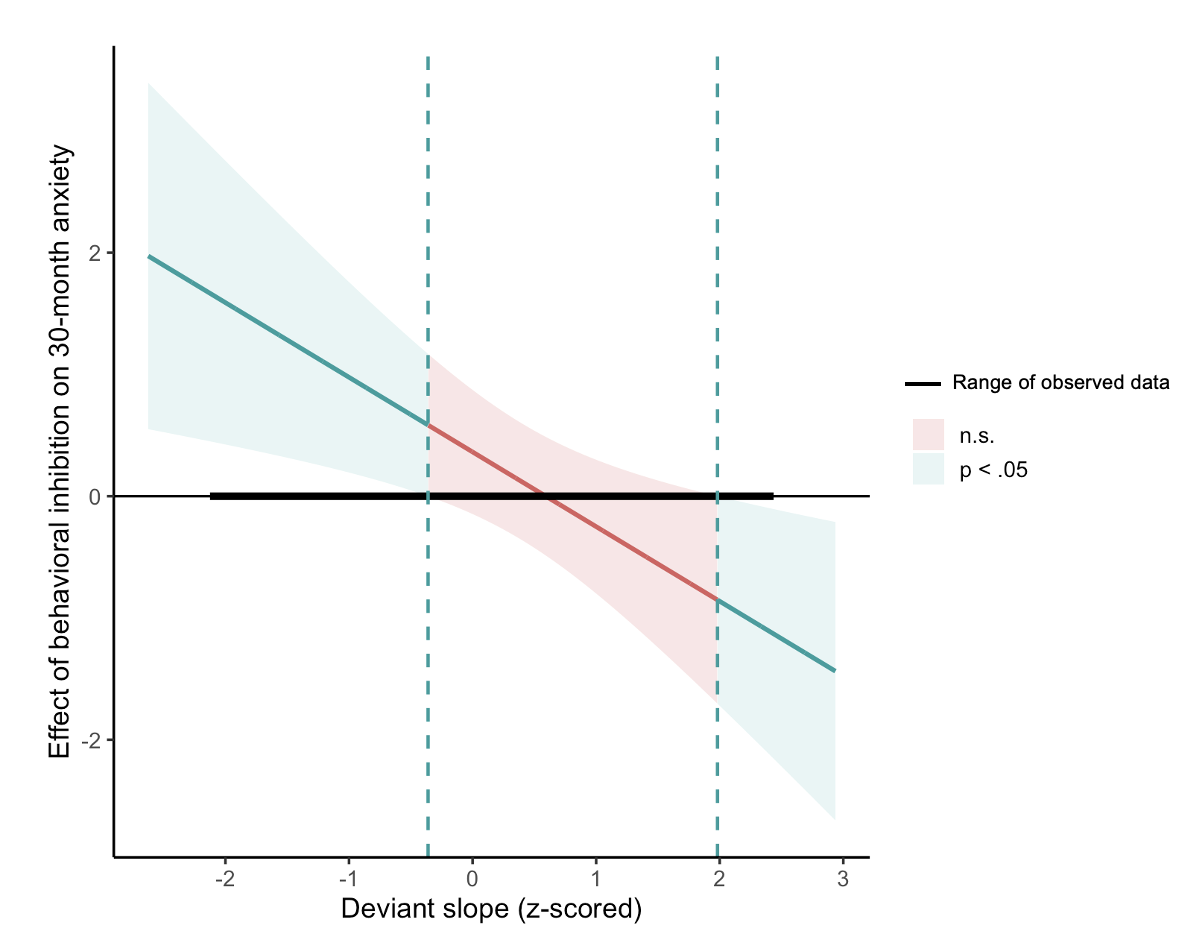

**Figure S4.** Regions of significance plot for sensitivity analysis. The blue and red shaded areas represent the levels of the moderator variable (slope) when the associations are statistically significant (*p <* .05) and non-significant (*p >* .05), respectively. Infants who exhibited a more negative slope (i.e., larger increase in prediction error over time) at 4 months are more likely to show a positive association between BI and anxiety, and those who exhibited a more positive slope (i.e., larger decrease in prediction error over time) at 4 months are more likely to show a negative association between NR and BI.

Overall, we partially replicated in the OIT cohort the prior findings in Xing et al. (2026), providing initial evidence of individual differences in the rate of changes in neural responses to deviance or the modulation of prediction error across time as potential mechanism underlying individual differences in MMR amplitudes in infancy. Interestingly, this effect emerged despite no group-level effect of time/block, indicating that the null average masked heterogeneity at the trial level. In our sample, some infants showed increasing and others decreasing neural responses to deviance over time which when aggregated canceled each other out. While the mechanisms underlying the directionality of the mismatch response in infancy still need further investigation, our results suggest that interrogating the trial-level patterns can inform how these average responses unfold over time. Our results show that infants with higher BI who show more negative slopes (which may index larger increase in prediction errors over time), are more likely to develop anxiety. Interestingly, a potential resilience effect also emerged, such that infants with higher BI but more positive slopes, which may index larger reductions in prediction error over time, may be less likely to develop anxiety. Although the resilience effect was not observed in the focal model reported in the Main Text, this aligned with the resilience effect found in Xing et al. (2026), and highlighted the potential that alterations in early sensory processing facilitating saliency detection could underlie anxiety risk and resilience pathway as early as 4 months. While we were not able to replicate Xing et al. (2026) in finding such effects on the pathway from NR to BI, the differences in NR distribution, developmental timepoints at which measures were administered, number of blocks in the EEG paradigm, may all have contributed to these discrepant findings, warranting additional replication.

**Sensitivity analysis: replication of the focal model using alternative NR calculations.**

We conducted sensitivity analyses to test the robustness of the findings to different ways of assessing and calculating NR. The focal model in the main text was replicated by examining the following two alternative NR variables: (1) the negative affect subscale score derived from the behavioral coding system reported in the main text; and (2) the fearful temperament assessed using a parent-report questionnaire, Infant Behavior Questionnaire (IBQ; (Gartstein & Rothbart, 2003).

***Parent Report of Infant Fearful Temperament*** At the 4-month visit, mothers completed the IBQ, a widely used caregiver-report measure designed to assess infant temperament. Caregivers report the frequency of specific infant behaviors across common caregiving situations during the previous week, using a 7-point Likert scale ranging from *never* to *always*. The questionnaire assesses multiple dimensions of early temperament, including activity level, distress to limitations, fear, duration of orienting, smiling and laughter, and soothability. Subscale scores were calculated by averaging the items corresponding to each dimension, with higher scores indicating greater expression of the trait. In the current study, the IBQ *Fear* and *Distress to Limitations* subscales were used to index early temperamental reactivity (*α* = .84 for the full questionnaire; *α* = .70 and .51 for *Fear* and *Distress to Limitations* subscales, respectively). A negative reactivity (NR) composite was calculated by multiplying standardized scores of the *Fear* and *Distress to Limitations subscales* following Filippi et al. (Filippi et al., 2024).

Overall, the MMR moderation effect on the NR–BI association didn’t remain significant across the alternative operationalizations of NR (*p*s > .48), demonstrating the specificity of the effect to the classic measure of NR (i.e., the product of observed negative affect and motor reactivity). The NR measure that incorporates both motor and affective responses may provide a more comprehensive index of Kagan’s classic temperament measure than the negative affect subscale alone. Additionally, the behaviorally coded NR measures may capture more nuanced differences in early temperament compared to parent-report questionnaires (i.e., IBQ) which exhibited low internal consistency on subscales relevant to temperament in the current sample.

**Sensitivity analysis: replication of the focal model with alternative BI calculations.**

We also replicated the focal SEM model using alternative measures of behavioral inhibition (BI) at 14 months, including the social fear subscale of the Toddler Behavior Assessment Questionnaire (TBAQ; Goldsmith, 1996) and a composite score averaging standardized observed BI and TBAQ–social fear scores following prior literature (e.g., Abend et al., 2018; Filippi et al., 2020; Xing et al., 2026). At the 14-month visit, mothers completed the TBAQ, a widely used caregiver-report measure designed to assess individual differences in toddler temperament across everyday contexts. The questionnaire asks parents to rate the frequency of specific behaviors their child displays in common situations on a 7-point Likert scale, with higher scores indicating greater expression of the temperament trait. In the current study, the *Social Fear* subscale was used as a proxy for behavioral inhibition (BI), as it captures toddlers’ wariness or distress in response to unfamiliar people and social situations (*α* = .90 for the full questionnaire; *α* = .64 for the *Social Fear* subscale). A BI composite score was further computed by averaging the z-scored observed BI and TBAQ-social fear subscale scores at 14 months.

The MMR moderation effects were not replicated when the TBAQ Social Fear subscale score or BI composite score was used (*p*s > .09), demonstrating the specificity of the effects to the measure of observed BI. The lack of significance may be attributed to that the behaviorally coded BI measure at 14 months may more sensitively index individual differences in early temperament compared to parent-report questionnaires (i.e., TBAQ), which also demonstrated low internal consistency on *Social Fear* subscale in the current sample.

**Sensitivity analysis: replication of the focal model with ERPs extracted at Fz.**

Given that prior work has extracted neural responses to unexpected stimuli at Fz (e.g., Kanel et al., 2025; Marshall et al., 2009; Xing et al., 2026), we additionally extracted MMR and P3 neural responses at Fz for sensitivity analysis. See Figure S4 below for the waveform and topo plots. Consistent with our visual inspection of the waveforms and topographies, MMR and P3 responses extracted at Fz were smaller than those extracted at F3/F4 (*p =* .097 and *p <* .001, respectively), supporting the selection of F3/F4 as the electrode clusters for analysis.

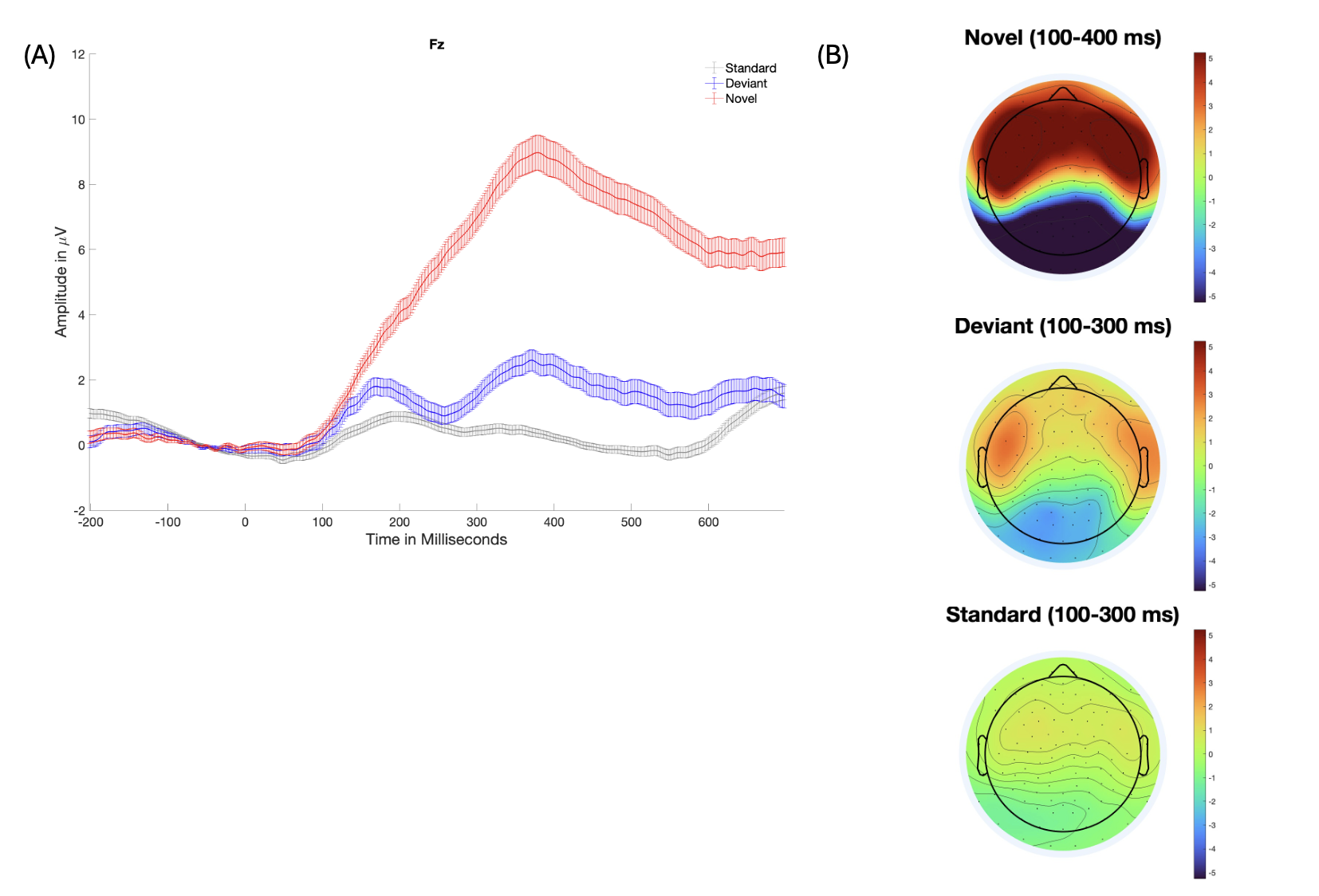

**Figure S5.** (A) ERP waveforms for standard, deviant and novel conditions at Fz electrode cluster; (B) The scalp topographic maps of neural responses in each condition.

For replication of the focal SEM model, the amplitude of the MMRs at 4 months extracted at Fz did not significantly moderate the association between NR at 4 months and BI at 14 months (*β* = -.08, *p* = .64), but significantly moderated the association between BI at 14 months and anxiety outcome at 30 months (*β* = -.64, *p* = .007). The MMR amplitude at 4 months also negatively correlated with BI at 14 months (*β* = -.26, *p* = .01). The anxiety outcomes across 30 months and 4-6 years were significantly associated (*β* = .54, *p* = .03). After removing non-significant paths, the SEM model revealed good model fit (χ^2^ (4)= .61, *p =* .96; CFI= 1; RMSEA= 0; SRMR = .06)(Hu & Bentler, 1999) (See Figure S5). Post-hoc probing analyses using model-implied conditional effects revealed that higher BI related to greater anxiety at 30 months, but only among infants with more negative MMRs at 4 months (*p <* .05, z-score range [-3.21, -0.87]). Infants with more positive MMRs at 4 months (z-score, range [-0.87, 2.56]) showed no significant associations between BI and anxiety (See Figure S7).

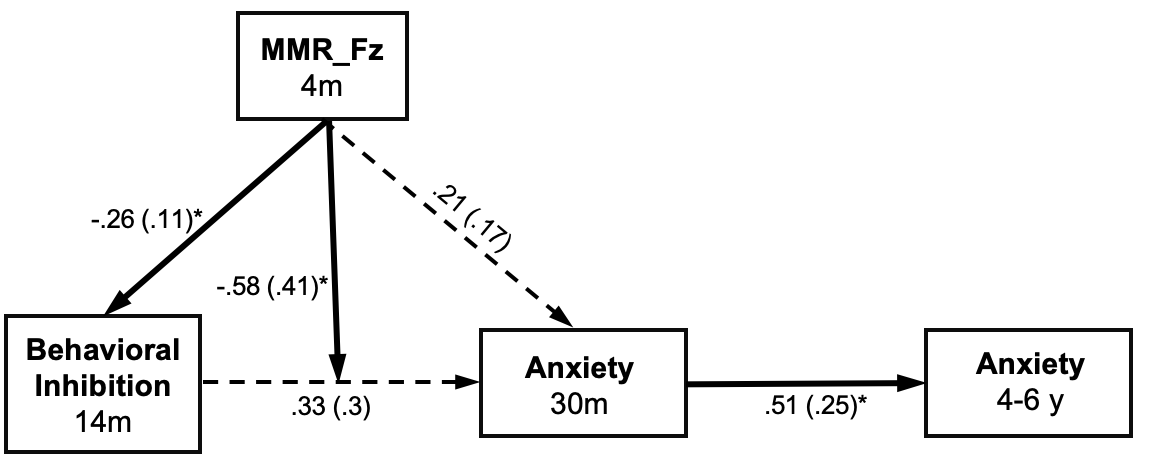

**Figure S6**. Sensitivity analysis with MMRs at Fz. Standardized parameter estimates are shown with standard error in parentheses. Statistically significant (*p* < .05) paths are indicated with solid lines and asterisks while non-significant paths are indicated with dashed lines. **p* < .05.

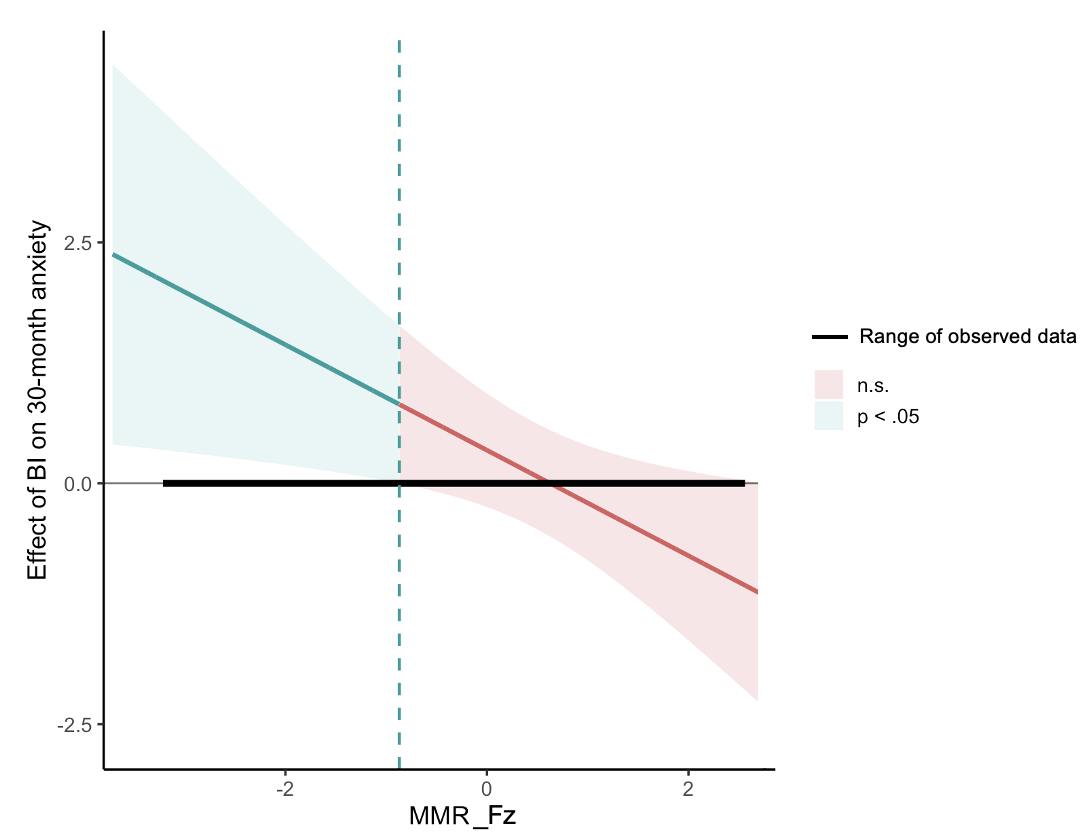

**Figure S7.** Regions of significance plots for the focal model. The blue and red shaded areas represent the levels of the moderator variable (MMRs) when the associations are statistically significant (*p* < .05) and non-significant (*p* > .05), respectively. Figure illustrates that infants with more negative MMRs at 4 months extracted at Fz show a positive association between behavioral inhibition and anxiety. In contrast, there is no significant association between BI and anxiety at more positive MMRs extracted at Fz.

| **Table S1A.** Correlations among variables included in the sensitivity analyses (left part of the matrix). | | | | | | |
| --- | --- | --- | --- | --- | --- | --- |
| Variables | | *n* | *M* | *SD* | 1 | 2 |
| **Negative Reactivity (NR)** | |  |  |  |  |  |
| 1. Multiplication of Negative Affect*Motor reactivity | | 130 | 0.32 | 1.22 |  |  |
| 2. IBQ_composite | | 111 | 0.36 | 1 | 0.2^*^ |  |
|  |  |  |  |  | [0.01, 0.37] |  |
| 3. Negative Affect | | 130 | 2.36 | 1.43 | 0.43^**^ | 0.18 |
|  |  |  |  |  | [0.27, 0.56] | [-0.008, 0.35] |
| **Behavioral Inhibition (BI)** | |  |  |  |  |  |
| 4. Observed BI | | 64 | -0.02 | 5.67 | 0.2 | 0.09 |
|  |  |  |  |  | [-0.05, 0.43] | [-0.16, 0.33] |
| 5. TBAQ_social fear | | 93 | 3.91 | 1.02 | 0.05 | 0.04 |
|  |  |  |  |  | [-0.16, 0.25] | [-0.17, 0.24] |
| 6. BI_composite | | 64 | -0.04 | 0.8 | 0.16 | 0.08 |
|  |  |  |  |  | [-0.09, 0.39] | [-0.17, 0.32] |
| 7. P3 | | 90 |  | 2.91 | 0.152 | -0.198 |
|  |  |  |  |  | [-0.06, 0.35] | [-0.41, 0.04] |
| 8. MMR | | 90 |  | 2.16 | -0.1 | -0.17 |
|  |  |  |  |  | [-0.30, 0.12] | [-0.39, 0.07] |
| 9. Attention Problems_30m | | 23 | 2.43 | 1.65 | 0.14 | -0.03 |
|  |  |  |  |  | [-0.29, 0.52] | [-0.44, 0.39] |
| 10. Anxiety_30m | | 23 | 2.09 | 1.98 | -0.14 | 0.4 |
|  |  |  |  |  | [-0.53, 0.28] | [-0.01, 0.7] |
| 11. Externalizing_30m | | 23 | 10.3 | 6.92 | -0.26 | -0.02 |
|  |  |  |  |  | [-0.61, 0.17] | [-0.43, 0.40] |
| 12. Attention Problems_4-6y | | 74 | 1.88 | 1.84 | 0.1 | -0.03 |
|  |  |  |  |  | [-0.13, 0.32] | [-0.26, 0.2] |
| 13. Anxiety_4-6y | | 74 | 2.8 | 2.61 | -0.17 | -0.15 |
|  |  |  |  |  | [-0.39, 0.06] | [-0.37, 0.09] |
| 14. Externalzing_4-6y | | 72 | 9.35 | 7.76 | -0.06 | -0.06 |
|  |  |  |  |  | [-0.28, 0.18] | [-0.29, 0.18] |
| *Note.* *M* and *SD* are used to represent mean and standard deviation, respectively. Values in square brackets indicate the 95% confidence interval for each correlation. The confidence interval is a plausible range of population correlations that could have caused the sample correlation. * indicates *p* < .05; ** indicates *p* < .01; *** indicates *p* < .001. | | | | | | |

| **Table S1B.** Correlations among variables included in the sensitivity analyses (middle part of the matrix). | | | | | | |
| --- | --- | --- | --- | --- | --- | --- |
| Variables | 3 | 4 | 5 | 6 | 7 | 8 |
| **Negative Reactivity (NR)** |  |  |  |  |  |  |
| 1. Multiplication of Negative Affect*Motor reactivity | 0.43^**^ | 0.2 | 0.05 | 0.16 | 0.153 | -0.109 |
|  | [0.27, 0.56] | [-0.05, 0.43] | [-0.16, 0.25] | [-0.10, 0.39] | [-0.06, 0.35] | [-0.31, 0.11] |
| 2. IBQ_composite | 0.18 | 0.1 | 0.04 | 0.08 | -0.19 | -0.19 |
|  | [-0.01, 0.36] | [-0.16, 0.33] | [-0.17, 0.24] | [-0.17, 0.32] | [-0.41, 0.04] | [-0.41, 0.04] |
| 3. Negative Affect |  | 0.19 | 0.19 | 0.14 | -0.21 | -0.19 |
|  |  | [-0.39, 0.17] | [-0.06, 0.42] | [-0.11, 0.38] | [-0.4, 0.01] | [-0.39, 0.02] |
| **Behavioral Inhibition (BI)** |  |  |  |  |  |  |
| 4. Observed BI | 0.19 |  | 0.46^**^ | 0.88^**^ | 0.07 | -0.118 |
|  | [-0.06, 0.42] |  | [0.25, 0.64] | [0.81, 0.92] | [-0.22, 0.35] | [-0.39, 0.17] |
| 5. TBAQ_social fear | -0.002 | 0.46^**^ |  | 0.83^**^ | -0.01 | 0.2 |
|  | [-0.21, 0.2] | [0.25, 0.64] |  | [0.74, 0.90] | [-0.25, 0.23] | [-0.05, 0.42] |
| 6. BI_composite | 0.14 | 0.88^**^ | 0.83^**^ |  | -0.02 | -0.03 |
|  | [-0.11, 0.38] | [0.81, 0.92] | [0.74, 0.90] |  | [-0.23, 0.27] | [-0.31, 0.26] |
| 7. P3 | -0.17 | 0.04 | -0.1 | -0.08 |  | 0.31^**^ |
|  | [-0.37, 0.05] | [-0.25, 0.33] | [-0.33, 0.15] | [-0.36, 0.21] |  | [0.11, 0.49] |
| 8. MMR | -0.26^*^ | -0.3^*^ | 0.15 | -0.19 | 0.11 |  |
|  | [-0.44, -0.05] | [-0.54, -0.02] | [-0.1, 0.37] | [-0.45, 0.1] | [-0.1, 0.31] |  |
| 9. Attention Problems_30m | -0.11 | 0.1 | 0.16 | 0.19 | -0.02 | 0.17 |
|  | [-0.50, 0.31] | [-0.46, 0.6] | [-0.27, 0.54] | [-0.38, 0.65] | [-0.46, 0.43] | [-0.29, 0.57] |
| 10. Anxiety_30m | -0.09 | 0.41 | 0.4 | 0.35 | -0.1 | 0.3 |
|  | [-0.48, 0.34] | [-0.15, 0.77] | [-0.02, 0.70] | [-0.22, 0.74] | [-0.52, 0.36] | [-0.16, 0.66] |
| 11. Externalizing_30m | 0.14 | 0.47 | 0.17 | 0.47 | -0.01 | 0.24 |
|  | [-0.29, 0.52] | [-0.08, 0.80] | [-0.26, 0.54] | [-0.08, 0.80] | [-0.45, 0.44] | [-0.22, 0.62] |
| 12. Attention Problems_4-6y | -0.02 | -0.02 | 0.02 | -0.02 | -0.13 | -0.26 |
|  | [-0.25, 0.21] | [-0.32, 0.28] | [-0.23, 0.26] | [-0.32, 0.28] | [-0.4, 0.18] | [-0.51, 0.04] |
| 13. Anxiety_4-6y | 0 | 0.13 | 0.15 | 0.12 | -0.06 | 0.11 |
|  | [-0.23, 0.23] | [-0.18, 0.41] | [-0.10, 0.38] | [-0.18, 0.40] | [-0.34, 0.24] | [-0.19, 0.39] |
| 14. Externalzing_4-6y | 0.05 | 0.15 | 0.15 | 0.21 | -0.26 | -0.25 |
|  | [-0.18, 0.28] | [-0.16, 0.43] | [-0.1, 0.38] | [-0.10, 0.47] | [-0.52, 0.03] | [-0.51, 0.04] |
| *Note.* *M* and *SD* are used to represent mean and standard deviation, respectively. Values in square brackets indicate the 95% confidence interval for each correlation. The confidence interval is a plausible range of population correlations that could have caused the sample correlation. * indicates *p* < .05; ** indicates *p* < .01; *** indicates *p* < .001. | | | | | | |

| **Table S1C.** Correlations among variables included in the sensitivity analyses (right part of the matrix). | | | | | |
| --- | --- | --- | --- | --- | --- |
| Variables | 9 | 10 | 11 | 12 | 13 |
| **Negative Reactivity (NR)** |  |  |  |  |  |
| 1. Multiplication of Negative Affect*Motor reactivity | 0.14 | -0.16 | -0.26 | 0.1 | -0.17 |
|  | [-0.29, 0.52] | [-0.53, 0.29] | [-0.61, 0.17] | [-0.13, 0.32] | [-0.39, 0.06] |
| 2. IBQ_composite | -0.03 | 0.4 | -0.02 | -0.03 | -0.15 |
|  | [-0.44, 0.39] | [-0.01, 0.70] | [-0.43, 0.40] | [-0.26, 0.20] | [-0.37, 0.09] |
| 3. Negative Affect | -0.11 | -0.09 | 0.14 | -0.02 | -0.001 |
|  | [-0.50, 0.31] | [-0.48, 0.34] | [-0.29, 0.52] | [-0.25, 0.21] | [-0.23, 0.23] |
| **Behavioral Inhibition (BI)** |  |  |  |  |  |
| 4. Observed BI | 0.1 | 0.41 | 0.47 | -0.02 | 0.13 |
|  | [-0.46, 0.60] | [-0.15, 0.77] | [-0.08, 0.80] | [-0.32, 0.28] | [-0.18, 0.41] |
| 5. TBAQ_social fear | 0.16 | 0.4 | 0.17 | 0.02 | 0.15 |
|  | [-0.27, 0.54] | [-0.02, 0.70] | [-0.23, 0.26] | [-0.23. 0.26] | [-0.10, 0.38] |
| 6. BI_composite | 0.19 | 0.35 | 0.48 | -0.02 | 0.12 |
|  | [-0.38, 0.65] | [-0.22, 0.77] | [-0.08, 0.80] | [-0.32, 0.28] | [-0.18, 0.40] |
| 7. P3 | 0.06 | -0.12 | -0.14 | -0.05 | -0.01 |
|  | [-0.39, 0.49] | [-0.54, 0.34] | [-0.55, 0.32] | [-0.34, 0.24] | [-0.31, 0.28] |
| 8. MMR | -0.12 | 0.25 | 0.22 | -0.23 | 0.18 |
|  | [-0.53, 0.34] | [-0.22, 0.62] | [-0.25, 0.60] | [-0.54, 0.001] | [-0.12, 0.45] |
| 9. Attention Problems_30m |  | 0.55^**^ | 0.48^*^ | 0.41 | 0.08 |
|  |  | [0.17, 0.78] | [0.09, 0.75] | [-0.4, 0.72] | [-0.37, 0.51] |
| 10. Anxiety_30m | 0.55^**^ |  | 0.68^**^ | 0.1 | 0.41 |
|  | [0.17, 0.78] |  | [0.35, 0.85] | [-0.36, 0.52] | [-0.04, 0.72] |
| 11. Externalizing_30m | 0.41^*^ | 0.87^**^ |  | -0.16 | 0.50^*^ |
|  | [-0.04, 0.72] | [-0.37, 0.51] |  | [-0.57, 0.30] | [0.07, 0.77] |
| 12. Attention Problems_4-6y | 0.41 | 0.1 | -0.16 |  | 0.49^*^ |
|  | [-0.04, 0.72] | [-0.34, 0.52] | [-0.57, 0.30] |  | [0.29, 0.65] |
| 13. Anxiety_4-6y | 0.08 | 0.41 | 0.50^*^ | 0.32^**^ |  |
|  | [-0.37, 0,51] | [-0.04, 0.72] | [0.07, 0.77] | [0.10, 0.51] |  |
| 14. Externalzing_4-6y | -0.02 | 0.09 | 0.16 | 0.65^**^ | 0.49^**^ |
|  | [-0.46, 0.42] | [-0.37, 0.51] | [-0.31, 0.56] | [0.49, 0.77] | [0.29, 0.65] |
| *Note.* *M* and *SD* are used to represent mean and standard deviation, respectively. Values in square brackets indicate the 95% confidence interval for each correlation. The confidence interval is a plausible range of population correlations that could have caused the sample correlation. * indicates *p* < .05; ** indicates *p* < .01; *** indicates *p* < .001. | | | | | |
